# Non-Participation Undermines Mass Drug Administration Effectiveness and Sustains Urogenital Schistosomiasis Transmission in an Endemic Community in Ghana

**DOI:** 10.64898/2026.08.03.26359546

**Authors:** Emmanuel Mensah, Alberta Serwah Anning, Emmanuella Akumeniwaa Nkrumah, Dorcas Sekyi, Suzzana Dickson Buabeng, Emmanuel Adu Sarpong, Nicholas Donkor, Frank Agyei, Godson Sodzah, Herbert Adjei, Gloria Hagan, Michael Asare, Prince Anaglatey, George Ghartey-Kwansah

**Author notes:** **Corresponding Author**: George Ghartey-Kwansah (PhD), **Email address**; Department of Biomedical Sciences, School of Allied Health Sciences, College of Health and Allied Sciences, University of Cape Coast, Cape Coast, Ghana.

## Abstract

Non-participation in MDA undermines schistosomiasis control and sustains local transmission hotspots in Ghana despite national progress. Schistosomiasis, a neglected tropical disease caused by parasitic flatworms of the genus *Schistosoma*, remains prevalent in sub-Saharan Africa due to poverty, poor access to clean water and sanitation, and low awareness. This study evaluated the impact of non-participation on MDA effectiveness in the Abee community, Central Region, while identifying barriers to uptake and strategies to improve community engagement. A cross-sectional study was conducted among 298 participants. Schistosomiasis prevalence was assessed by urine microscopy and PCR, and multivariate logistic regression was used to identify factors influencing infection and MDA participation (p ≤ 0.05). Results showed a prevalence of 17.1% by microscopy and 20.1% by PCR. Only 48.7% of participants had ever participated in MDA, with 31.5% never treated. Awareness of schistosomiasis was 60.4%. Multivariate analysis revealed that non-participation in MDA and lack of disease education significantly increased infection risk, while awareness strongly predicted MDA uptake. Microhaematuria and leukocyturia were detected in 6.7% and 8.4% of participants, respectively. These WASH gaps facilitate environmental contamination with *Schistosoma haematobium* eggs, sustaining transmission despite ongoing MDA efforts. In conclusion, low MDA participation driven by limited awareness and weak WASH infrastructure sustains focal transmission hotspots (17.1–20.1% prevalence) in Abee. Strengthening community engagement, targeted health education, improved MDA access, and integrated WASH interventions is essential to increase coverage and achieve the WHO 2030 elimination targets.

**Author Summary:** Schistosomiasis, also known as bilharzia, is a major neglected tropical disease that affects millions of people, particularly in sub-Saharan Africa. Although Ghana has made progress through mass drug administration (MDA) with praziquantel, the disease continues to persist in some communities. In this study, investigation was done to determine why schistosomiasis persists in the Abee community in Ghana’s Central Region. 298 participants were recruited and tested their urine using both conventional microscopy and a more sensitive molecular (PCR) method. 17.1% tested positive by microscopy and 20.1% by PCR. Strikingly, only 48.7% of participants had ever received praziquantel during MDA campaigns, and nearly one-third (31.5%) had never been treated. People who had never participated in MDA and those with low awareness of the disease were significantly more likely to be infected. Poor sanitation practices were also widespread, with many residents sharing toilets or practicing open defecation. These conditions allow the parasite to continue circulating in the environment. Our findings show that low participation in treatment programmes, limited health education, and inadequate water, sanitation, and hygiene (WASH) infrastructure are the main reasons schistosomiasis transmission persists in this community. This study highlights that MDA alone is not sufficient. Achieving the WHO 2030 elimination targets will require stronger community engagement, improved health education, higher treatment coverage, and better sanitation services.

## Introduction

Schistosomiasis remains one of the most important neglected tropical diseases, imposing a substantial health and socioeconomic burden in many low- and middle-income countries [1]. The disease is caused by blood flukes of the genus *Schistosoma*, of which five species commonly infect humans [2]. Transmission occurs through contact with freshwater contaminated by cercariae released from infected intermediate host snails, making communities with inadequate access to safe water, sanitation, and hygiene particularly vulnerable [3, 4]. Although considerable progress has been made in reducing the global burden of schistosomiasis, an estimated hundreds of millions of people remain at risk of infection, with the overwhelming majority of cases occurring in sub-Saharan Africa [1].

Among the human schistosome species, *Schistosoma haematobium* is the principal cause of urogenital schistosomiasis and is widely distributed across Africa and parts of the Middle East. Chronic infection is associated with haematuria, urinary tract pathology, renal impairment, infertility, and an increased risk of squamous cell carcinoma of the bladder. Consequently, urogenital schistosomiasis continues to represent a significant public health challenge, particularly in underserved rural communities where dependence on infested freshwater bodies for domestic, occupational, and recreational activities sustains transmission [1, 5].

Ghana remains endemic for both *S. haematobium* and *S. mansoni*, despite several decades of national control efforts. Preventive chemotherapy using praziquantel has substantially reduced the prevalence and intensity of infection in many endemic districts; however, transmission persists in numerous focal communities [6–8]. National surveys have demonstrated marked reductions in overall prevalence, yet localized hotspots continue to record moderate to high infection levels, indicating that transmission has not been interrupted uniformly across the country [5, 9–13]. These observations suggest that additional epidemiological and programmatic factors continue to limit progress towards elimination.

The cornerstone of schistosomiasis control is preventive chemotherapy with praziquantel, which has been recommended by the World Health Organization (WHO) for more than three decades. Following the adoption of World Health Assembly Resolution 54.19, regular mass drug administration (MDA) became the principal strategy for reducing schistosomiasis-related morbidity and interrupting transmission in endemic settings [9]. Administration of praziquantel at the recommended dose of 40 mg/kg body weight effectively reduces parasite burden, limits disease progression, and decreases environmental contamination by reducing egg excretion from infected individuals [10]. Nevertheless, chemotherapy alone cannot achieve elimination unless high treatment uptake is consistently maintained over successive rounds of MDA.

The effectiveness of MDA depends not only on the availability and distribution of praziquantel but also on the extent to which eligible individuals actually participate in treatment. Programme coverage generally refers to the proportion of the target population that receives treatment, whereas treatment compliance describes the proportion of individuals who ingest the medication after receiving it [14, 15]. These indicators are related but distinct, and both influence programme effectiveness. Individuals who repeatedly miss treatment may remain infected for prolonged periods, continue contaminating local water bodies, and sustain parasite transmission within their communities despite repeated MDA campaigns [16].

Although WHO has established ambitious coverage targets for schistosomiasis control, many endemic countries have struggled to consistently attain these goals. Global treatment coverage has historically lagged behind that achieved for several other neglected tropical diseases, limiting the impact of preventive chemotherapy programmes [17–19]. Beyond operational challenges, community-level factors, including absenteeism during drug distribution, misconceptions about treatment, fear of adverse drug reactions, inadequate health education, population mobility, and programme fatigue, can reduce participation in MDA and compromise programme outcomes [20–23]. Understanding these barriers is therefore essential for designing interventions that improve treatment uptake and accelerate progress towards elimination.

Despite the recognized importance of MDA participation, relatively few studies have comprehensively examined how non-participation influences the persistence of urogenital schistosomiasis in endemic Ghanaian communities after multiple treatment rounds. Identifying the characteristics of individuals who repeatedly miss treatment and determining their contribution to ongoing transmission are critical for optimizing control strategies and supporting the country’s elimination agenda.

Therefore, this study investigated the relationship between non-participation in mass drug administration and the persistence of urogenital schistosomiasis in an endemic community in Ghana. Specifically, the study assessed the determinants of non-participation and evaluated how missed treatment influences infection prevalence and the continued transmission of S. haematobium. The findings are expected to provide evidence to inform targeted interventions aimed at improving treatment uptake and strengthening national schistosomiasis elimination efforts.

## Methods

### Study design

This study employed a cross-sectional descriptive design using a mixed-methods approach to assess the effect of non-participation in mass drug administration (MDA) on schistosomiasis control in Abee, a community in the Komenda-Edina-Eguafo-Abirem (KEEA) Municipality of Ghana’s Central Region. Abee is situated along the Srowie River, a perennial freshwater body that supports the transmission of *Schistosoma haematobium* by providing suitable habitats for intermediate host snails. The river is extensively used for domestic, agricultural, and recreational purposes, resulting in frequent human–water contact. Most residents are indigenous farmers whose occupational and daily water-related activities increase their risk of exposure to schistosome-infested water and subsequent infection [24]. The study integrated quantitative parasitological diagnostics and structured questionnaires with qualitative data to capture both biomedical outcomes and the sociocultural factors influencing treatment uptake, compliance, and programme performance. Data collection was conducted from 2 April to 25 June 2026.

### Sample size

The sample size was determined statistically as n = 298 based on n=Z2⋅p⋅(1−p)E2, a formula for estimating a population proportion in a cross-sectional study [25]. This was calculated using a confidence interval of 95% (z = 1.96), a margin of error of 0.05, and an average estimated prevalence of 20% from other studies. This yielded a base sample size of approximately 246. To account for potential non-response or incomplete data, an additional 20% was added (246 × 1.2 = 295.2), and the sample size was rounded up to 298 for practical implementation. This adjustment ensures sufficient data to achieve the study’s objectives while accommodating challenges in participant recruitment.

### Ethical considerations

This study was conducted in accordance with national and international ethical guidelines for research involving human participants, with particular consideration for children. Ethical approval was obtained from the Institutional Review Board of the University of Cape Coast (UCCIRB/EXT/2024/043). Written informed consent was obtained from parents or guardians before data collection, and child assent was also secured using age-appropriate language to ensure participants’ understanding of the study. Participation was entirely voluntary, and participants were informed of their right to withdraw at any time without penalty, as well as the potential risks and benefits of the study. Confidentiality and anonymity were strictly maintained; no personal identifiers were included in data sets or reports, and unique codes were used for all participants. Data were securely stored in password-protected electronic files and locked cabinets, accessible only to authorized research personnel, and were used exclusively for academic purposes.

### Questionnaires

Structured questionnaires were used to collect data from the participants on the demographics, MDA participation, compliance, coverage, and access to water.

### Experimental Protocols for Sample Collection and Handling

Urine samples were collected from participants using standard parasitological procedures. Sterile, wide-mouth, leak-proof plastic containers were provided for urine collection. Participants received clear instructions to ensure proper sample collection and minimize contamination. Mid-way urine samples were collected to coincide with peak egg excretion of *Schistosoma haematobium.* All samples were labeled with unique identification codes and transported to the laboratory in ice-filled insulated containers immediately after collection. Upon arrival at the laboratory, samples were processed immediately, with no preservatives or transport media used.

### Urine tests and examination

A macroscopic examination was conducted to assess appearance and color. Further biochemical tests were conducted on the urine using the dipstick method to check parameters such as blood, leukocytes, protein, and other analytes. Microscopic urine examination was done using the urine sedimentation method. The urine sample was placed in a 15ml centrifuge tube and centrifuged for about 5 minutes. The supernatant was discarded, and a drop of the sediment was placed on a glass slide. A cover-slip was placed over it, and the slide was examined for *S. haematobium* eggs using the light microscope, as previously done [26].

### DNA extraction and Polymerase chain reaction

DNA was extracted from urine samples using the Ezup Urine/Saliva DNA extraction kit. DNA extraction, elution, and purification were carried out according to the manufacturer’s recommendations. The DNA extracts were amplified using conventional polymerase chain reaction (PCR) targeting a trematode-specific marker (18S_Digenia_F: CAGCTATGGTTCCTTAGATCRTA & 18S_Digenia_R: TATTTTTCGTCACTACCTCCCCGT), followed by a *Schistosoma* genus-specific marker (ITS2_Schisto_F: GGAAACCAATGTATGGGATTATTG & ITS2_Schisto_R: ATTAAGCCACGACTCGAGCA)[27, 28]. Each PCR was performed in a total reaction volume of 20 µL, comprising 5 µL of the DNA extract, 10 µL of Apex-HF HS DNA Polymerase FS Master Mix, 0.5 µL each of reverse and forward primer, and 4 µL of double-distilled water. The reaction conditions included an activation step of 95 °C for 3 minutes, followed by 40 cycles of 94 °C for 30 seconds, 58.5 °C for 45 seconds, and 72 °C for 45 seconds, and a final extension at 72 °C for 10 minutes. Amplification products were visualized by agarose gel electrophoresis at 90V for 35 minutes.

### Data analyses

Data from structured questionnaires, parasitological diagnostics (microscopy and PCR), and urinalysis were entered, cleaned, and managed in Microsoft Excel 2021, then exported to SPSS version 20 (IBM Corp., Armonk, NY, USA) for statistical analysis. Descriptive statistics were used to summarize study variables, with categorical data presented as frequencies and percentages and continuous variables as means and standard deviations. Associations between socio-demographic factors, schistosomiasis infection, and MDA participation were examined using Chi-square tests for categorical variables and independent samples t-tests for group comparisons of infection intensity. Multivariate logistic regression was conducted to identify independent predictors of infection and MDA uptake, with adjusted odds ratios (AORs) and 95% confidence intervals (CIs) reported. All analyses were two-tailed, and p-values ≤ 0.05 were considered statistically significant.

## Results

Table 1 presents the baseline demographic, clinical, and parasitological characteristics of the study population from the Abee community. A total of 298 participants were enrolled through school-based and household-based recruitment and completed structured questionnaires and parasitological assessments. The table summarizes distributions by sex, recruitment setting (school vs. community), urine characteristics (colour and appearance), and schistosomiasis infection status. Females constituted 56.4% of the study population, and the majority of participants were recruited from the school-based cohort (78.9%). Urine samples were predominantly straw-coloured and clear. The prevalence of schistosomiasis was 17.1% (51/298) by microscopy and 20.1% (60/298) by PCR, indicating a higher detection yield with molecular diagnostics. Frequencies and proportions for all variables are reported in the table.

**Table 1:**
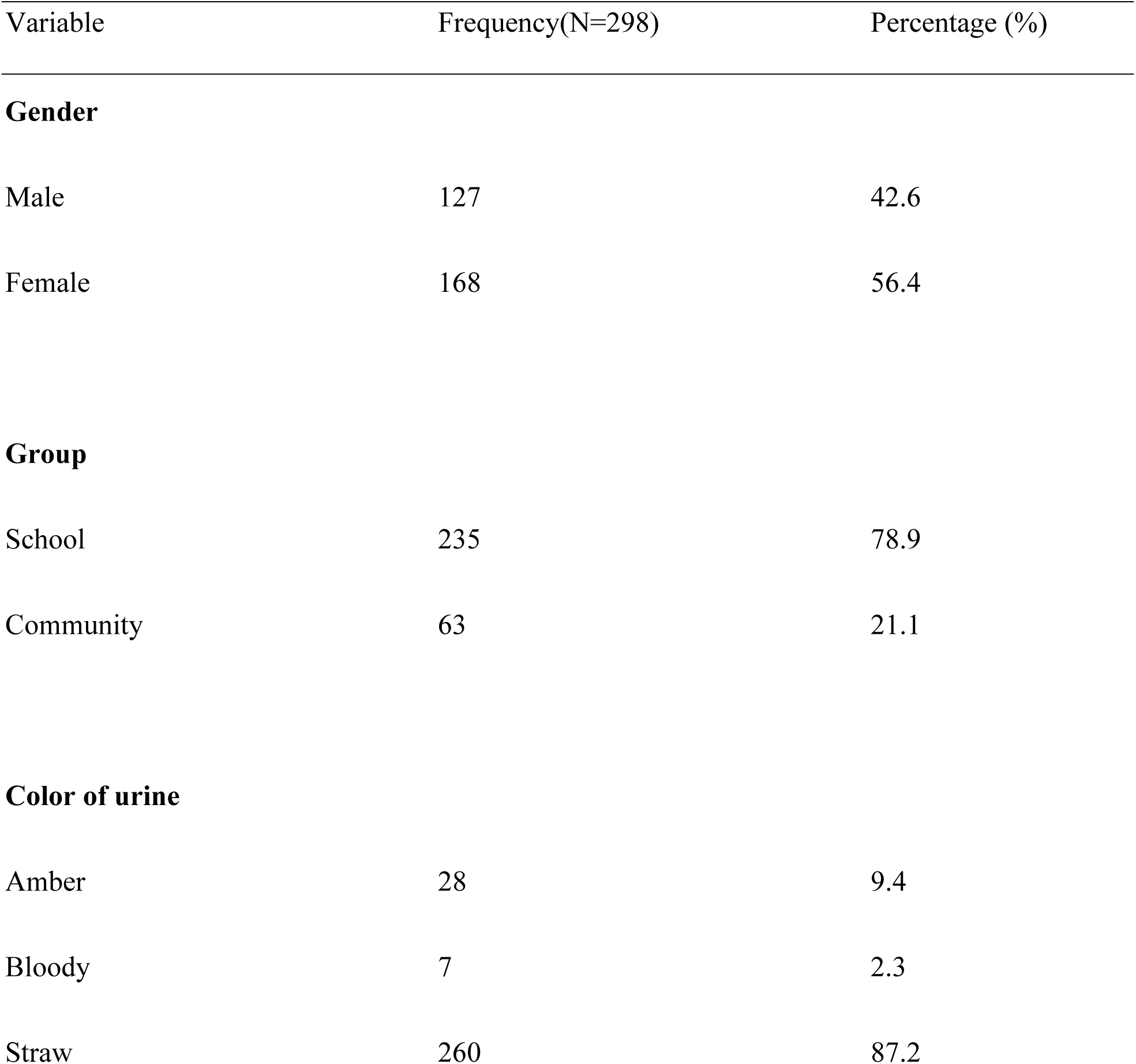
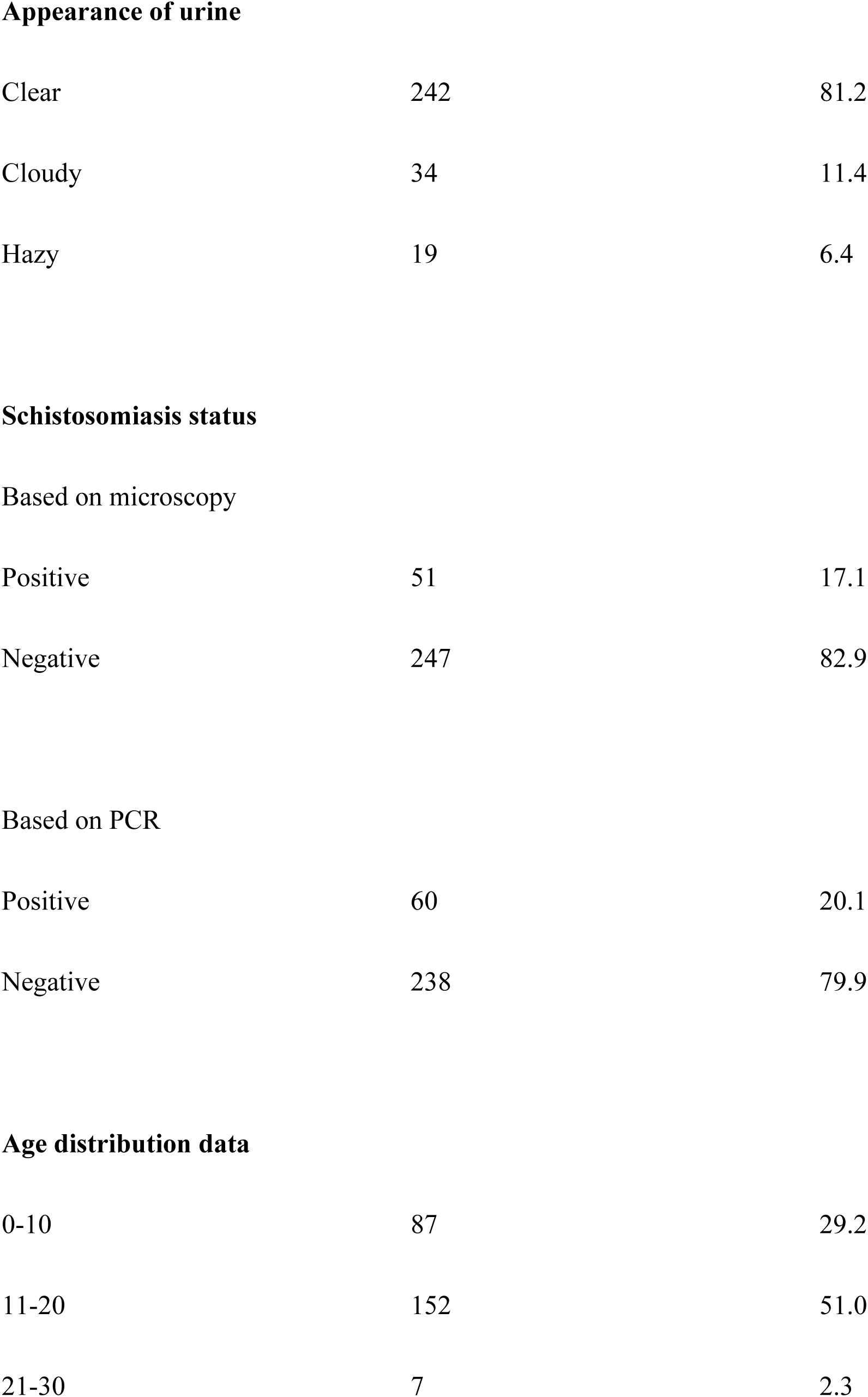
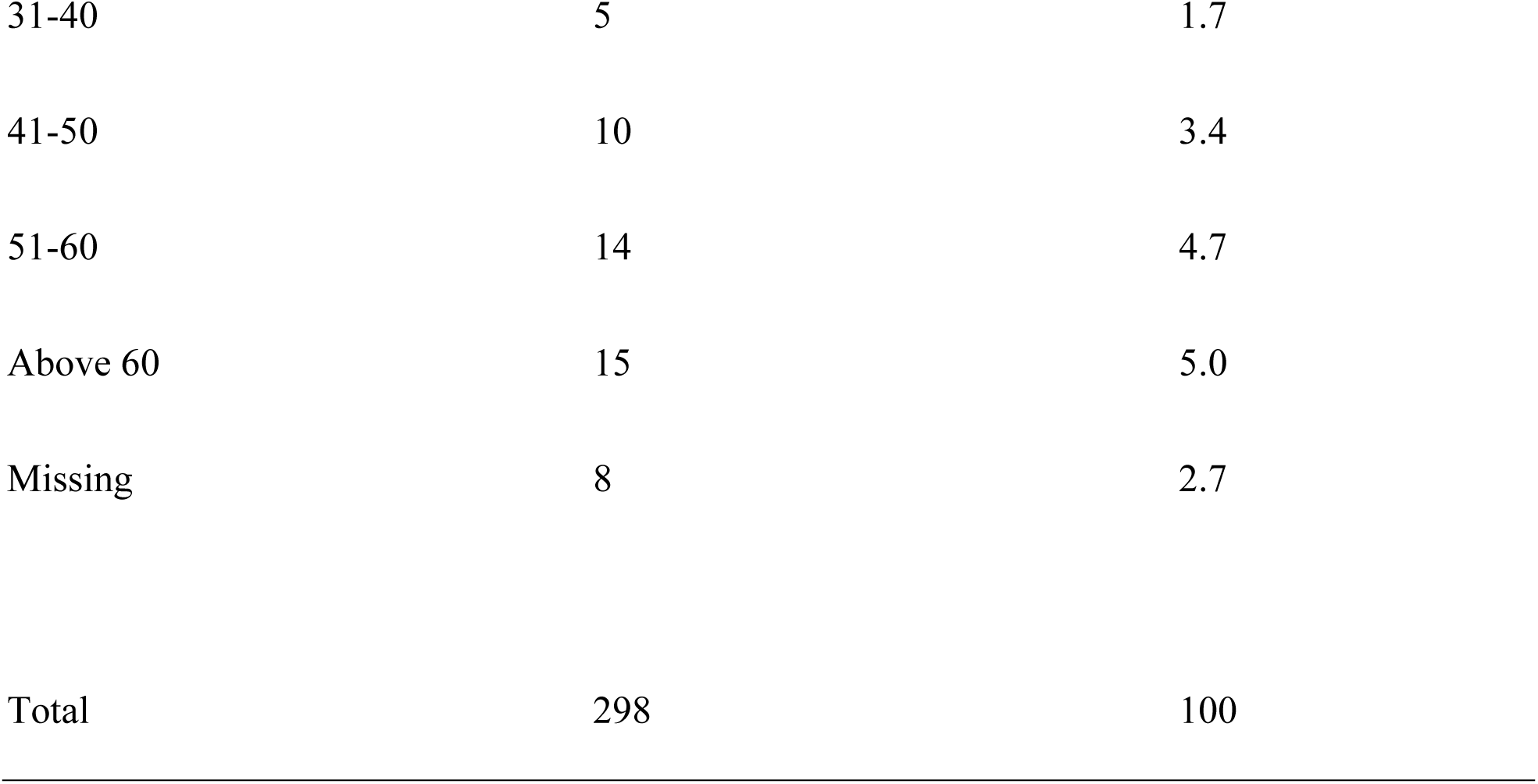
General characteristics of the study population.

| Variable | Frequency(N=298) | Percentage (%) |
| --- | --- | --- |
| **Gender** |  |  |
| Male | 127 | 42.6 |
| Female | 168 | 56.4 |
| **Group** |  |  |
| School | 235 | 78.9 |
| Community | 63 | 21.1 |
| **Color of urine** |  |  |
| Amber | 28 | 9.4 |
| Bloody | 7 | 2.3 |
| Straw | 260 | 87.2 |

|  |  |  |  |
| --- | --- | --- | --- |
| 243 | **Appearance of urine** |  |  |
| 244 | Clear | 242 | 81.2 |
| 245 | Cloudy | 34 | 11.4 |
| 246 | Hazy | 19 | 6.4 |
| 247 |  |  |  |
| 248 | **Schistosomiasis status** |  |  |
| 249 | Based on microscopy |  |  |
| 250 | Positive | 51 | 17.1 |
| 251 | Negative | 247 | 82.9 |
| 252 |  |  |  |
| 253 | Based on PCR |  |  |
| 254 | Positive | 60 | 20.1 |
| 255 | Negative | 238 | 79.9 |
| 256 |  |  |  |
| 257 | **Age distribution data** |  |  |
| 258 | 0-10 | 87 | 29.2 |
| 259 | 11-20 | 152 | 51.0 |
| 260 | 21-30 | 7 | 2.3 |
| 261 | 31-40 | 5 | 1.7 |
| 262 | 41-50 | 10 | 3.4 |
| 263 | 51-60 | 14 | 4.7 |
| 264 | Above 60 | 15 | 5.0 |
| 265 | Missing | 8 | 2.7 |
| 266 |  |  |  |
| 267 | Total | 298 | 100 |

In addition to baseline demographic and parasitological characteristics, Table 2 also presents patterns of MDA participation, awareness of schistosomiasis, and willingness to participate in treatment. Nearly half of the participants reported having ever received praziquantel (48.7%), while 60.4% indicated prior awareness of schistosomiasis.

**Table 2:**
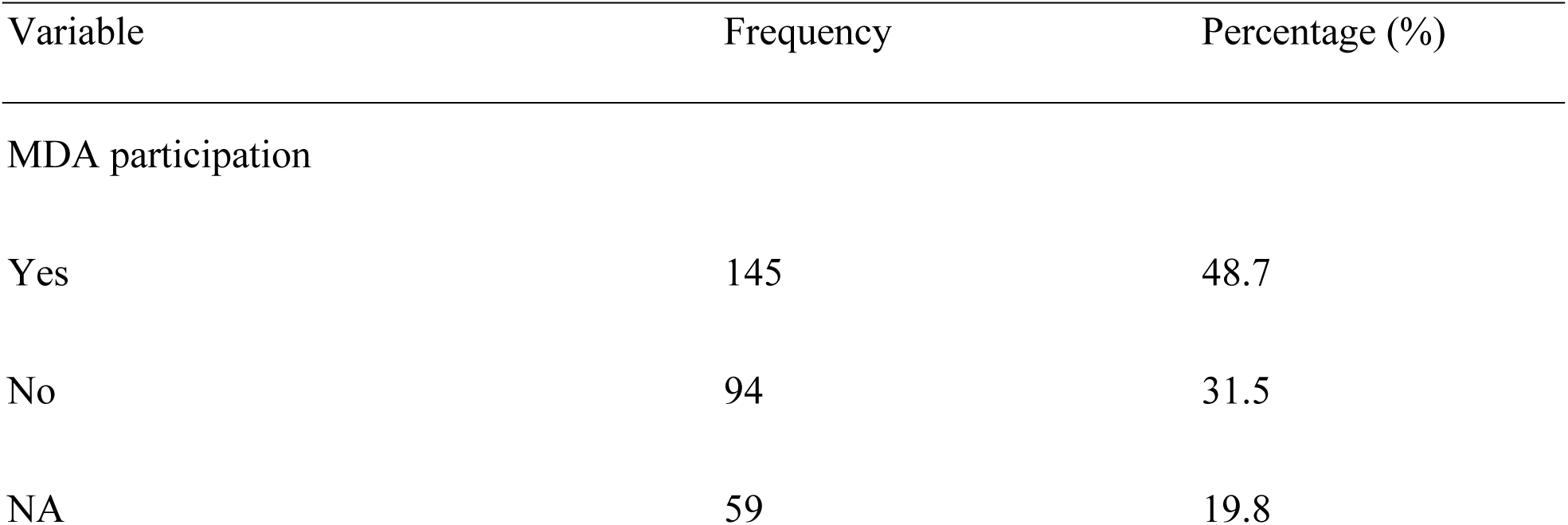
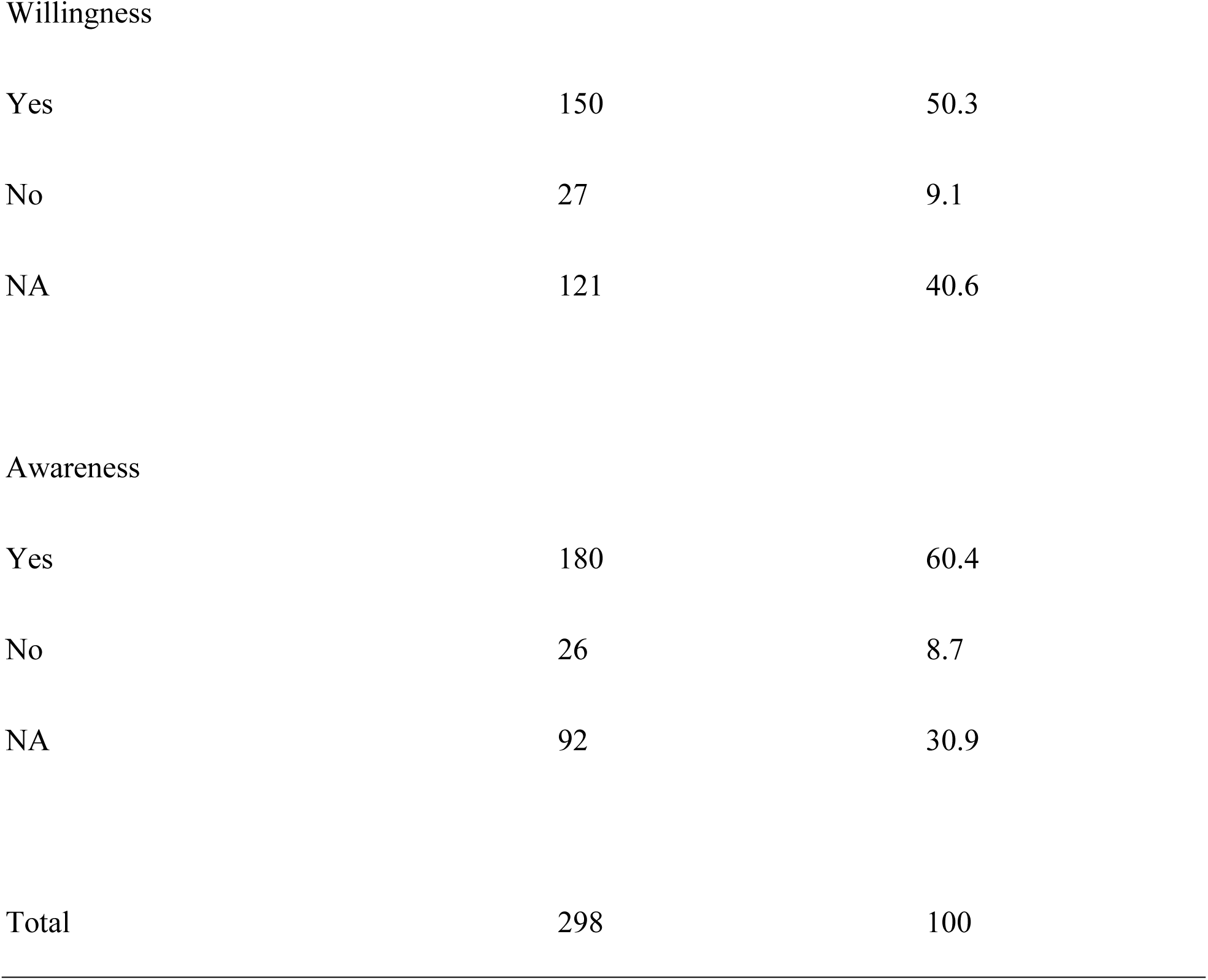
Mass drug administration participation, awareness, and willingness.

Table 3 summarizes sanitation practices in the Abee community. A substantial proportion of participants relied on suboptimal sanitation facilities, with 12.8% using public toilets, 11.4% pit latrines, 7.0% refuse dumps, and 2.7% open defecation (digging ground). Additionally, 43.3% reported sharing toilet facilities. These conditions facilitate environmental contamination with *Schistosoma haematobium* eggs, thereby maintaining the transmission cycle in water bodies despite ongoing MDA efforts.

**Table 3:** Analysis of variables associated with transmission dynamics.

| 300 | Variable | frequency | percentage (%) |
| --- | --- | --- | --- |
| 301 | **Type of toilet** |  |  |
| 302 | Public toilet | 38 | 12.8 |
| 303 | Pit latrine | 34 | 11.4 |
| 304 | WC | 71 | 23.8 |
| 305 | Refuse dump | 21 | 7.0 |
| 306 | Digging ground | 8 | 2.7 |
| 307 | Home latrine | 29 | 9.4 |
| 308 | All options | 38 | 12.8 |
| 309 |  |  |  |
| 310 |  |  |  |
| 311 | Sharing a toilet facility |  |  |
| 312 | Yes | 129 | 43.3 |
| 313 | No | 109 | 36.6 |

Urinalysis findings further indicated that the majority of participants (278/298) had an RBC count of 0, suggesting the absence of detectable haematuria in most individuals. Only a small proportion of participants exhibited elevated RBC counts (Table 4). Also, urinalysis revealed that the majority of participants (273/298) had a WBC count of 0, indicating the absence of detectable leukocyturia in most individuals. Only a small proportion of participants exhibited elevated WBC counts.

**Table 4:**
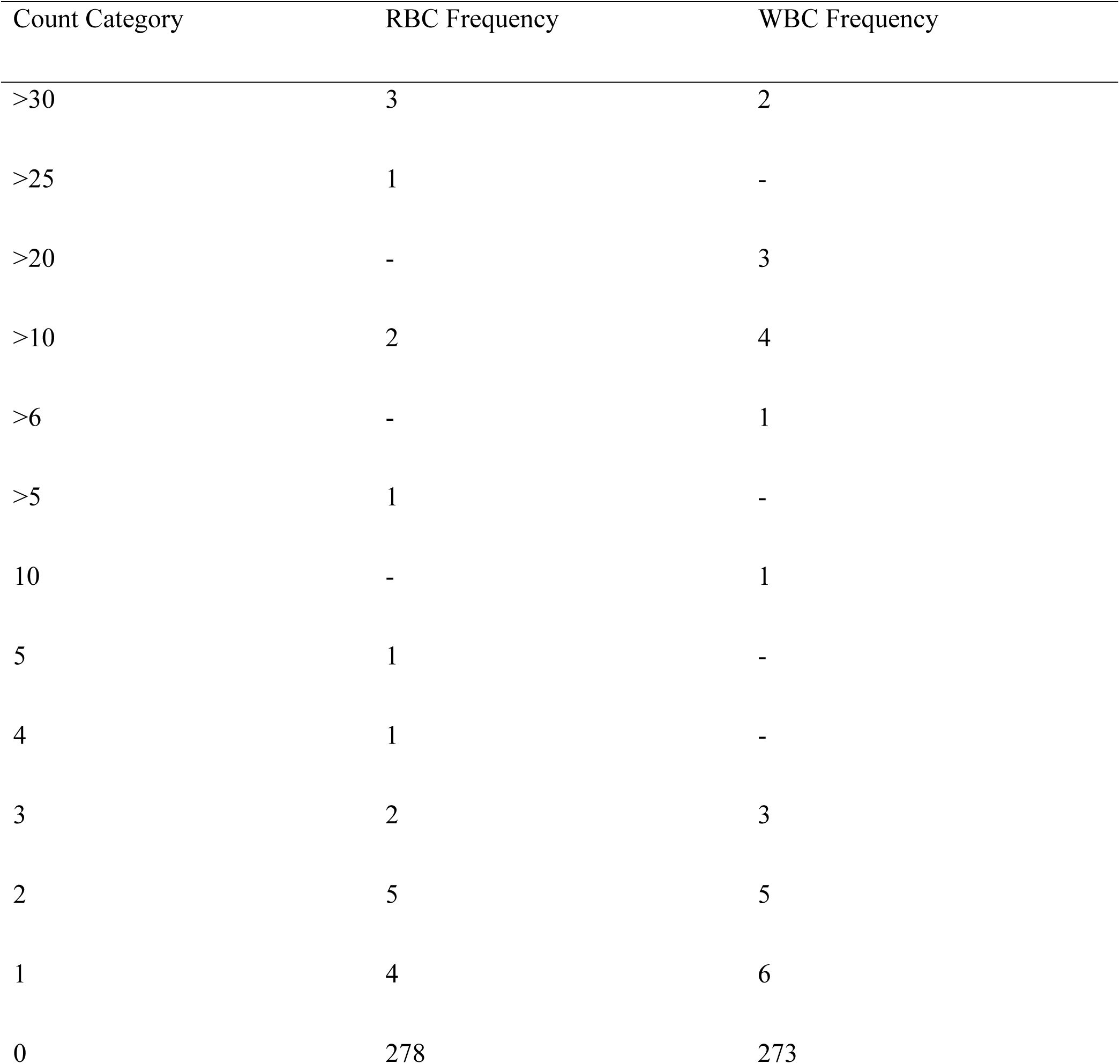
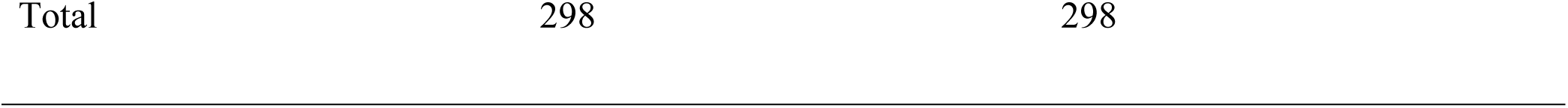
Urine Red blood cell (RBC) and white blood cell (WBC) counts.

Table 5 presents the multivariate analysis of factors associated with schistosomiasis infection. The model identifies key socio-demographic and exposure-related determinants, including knowledge of schistosomiasis, educational status, exposure to freshwater bodies, and history of treatment, as predictors of infection risk. Statistically significant associations were observed, notably the effect of education on schistosomiasis (p = 0.018), indicating its potential protective role against infection.

**Table 5:**
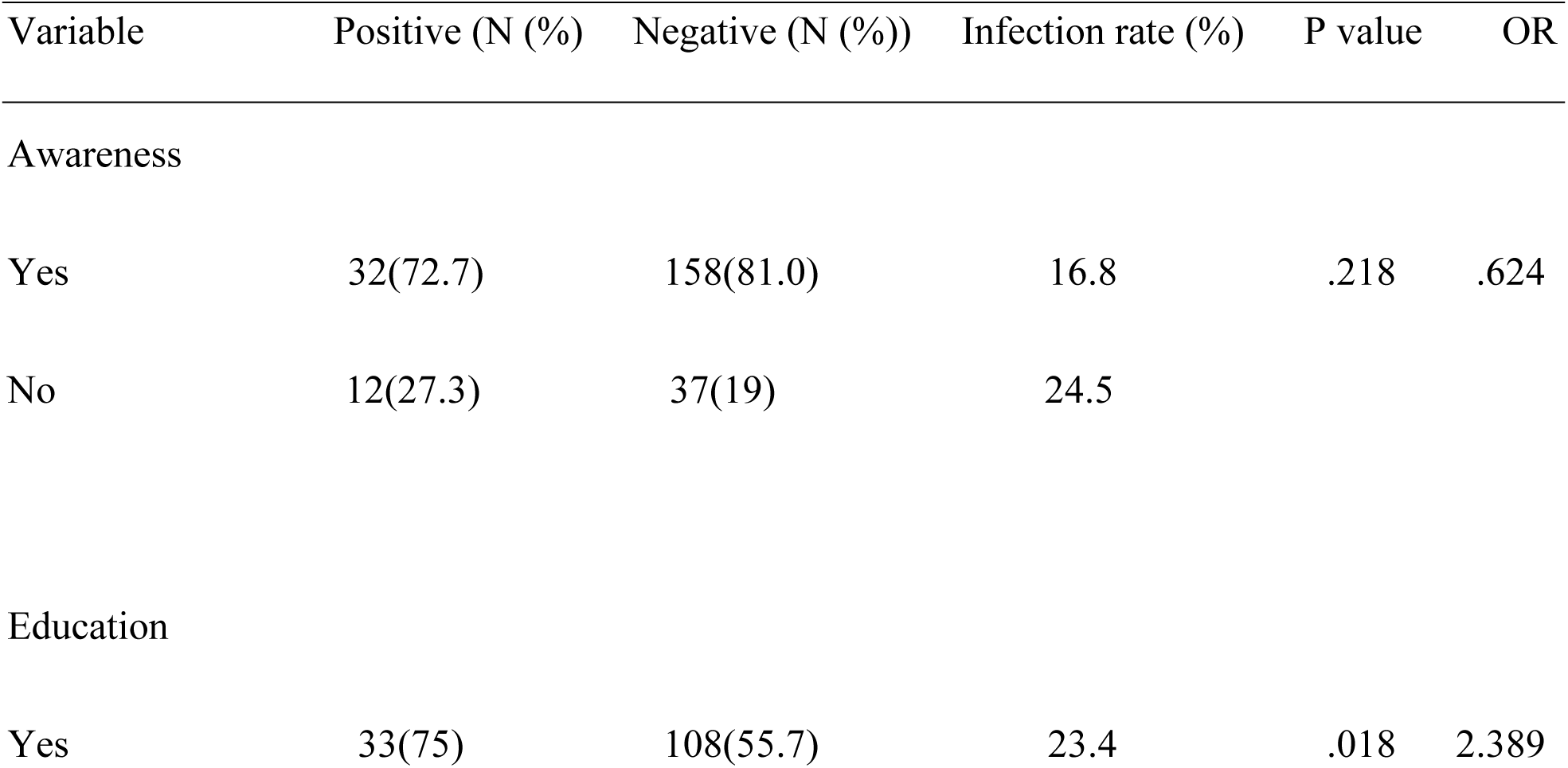
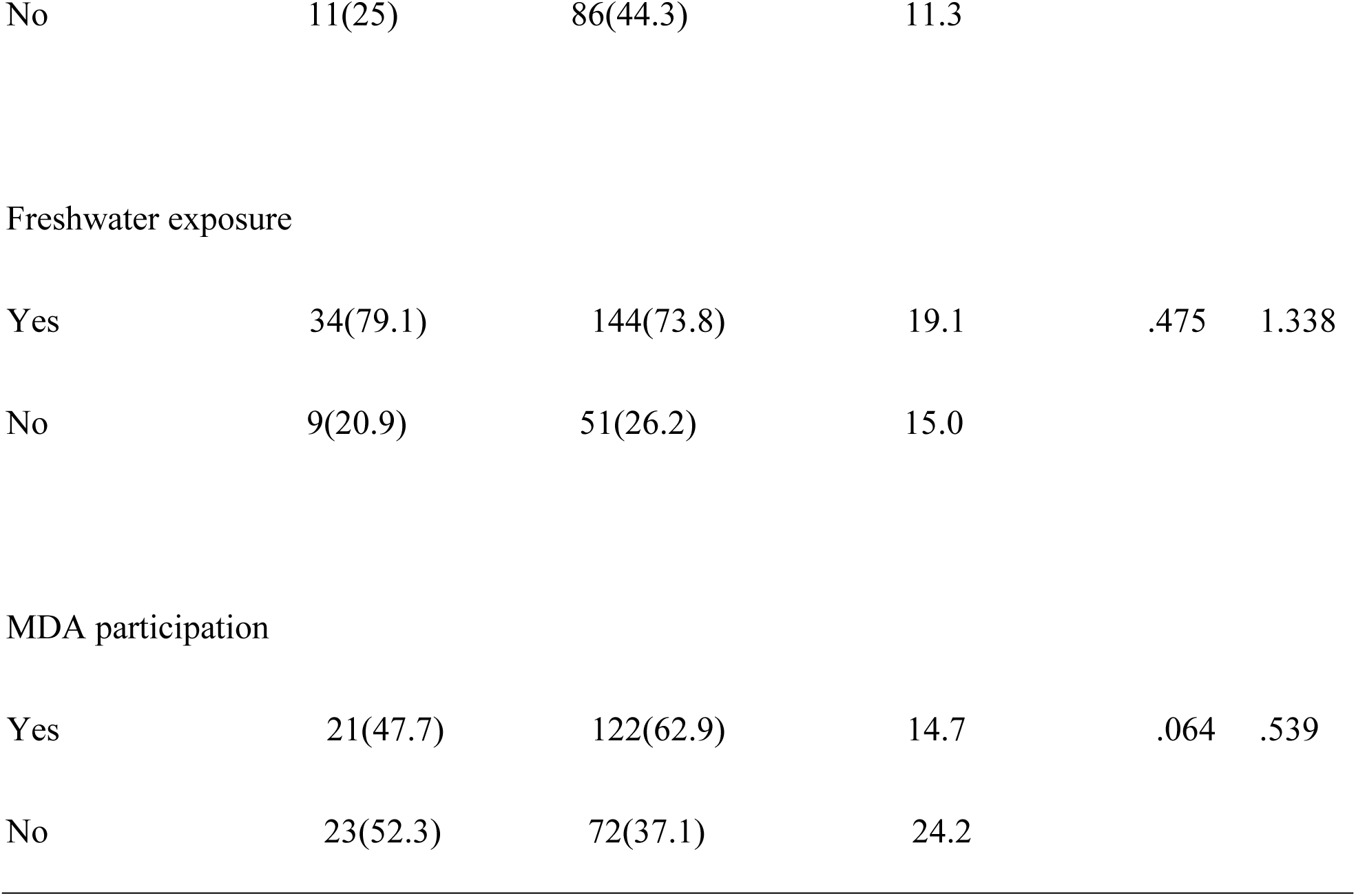
Multivariate analysis of variables associated with the frequency of schistosomiasis among study participants.

| Variable | Positive (N (%)) | Negative (N (%)) | Infection rate (%) | P value | OR |
| --- | --- | --- | --- | --- | --- |
| Awareness |  |  |  |  |  |
| Yes | 32(72.7) | 158(81.0) | 16.8 | .218 | .624 |
| No | 12(27.3) | 37(19) | 24.5 |  |  |
| Education |  |  |  |  |  |
| Yes | 33(75) | 108(55.7) | 23.4 | .018 | 2.389 |

|  |  |  |  |  |  |  |
| --- | --- | --- | --- | --- | --- | --- |
| 342 | No | 11(25) | 86(44.3) | 11.3 |  |  |
| 343 |  |  |  |  |  |  |
| 344 | Freshwater exposure |  |  |  |  |  |
| 345 | Yes | 34(79.1) | 144(73.8) | 19.1 | .475 | 1.338 |
| 346 | No | 9(20.9) | 51(26.2) | 15.0 |  |  |
| 347 |  |  |  |  |  |  |
| 348 | MDA participation |  |  |  |  |  |
| 349 | Yes | 21(47.7) | 122(62.9) | 14.7 | .064 | .539 |
| 350 | No | 23(52.3) | 72(37.1) | 24.2 |  |  |

Awareness of schistosomiasis emerged as a significant predictor of MDA participation (p = 0.004; OR = 2.570), indicating that individuals with prior knowledge of the disease were more than twice as likely to participate in MDA compared with those who lacked such awareness (Table 6).

**Table 6:** Multivariate analysis of variables associated with MDA effectiveness among study participants.

|  |  |  |  |  |  |
| --- | --- | --- | --- | --- | --- |
| 358 | Variable | MDA(Yes) | MDA(No) | P value | OR |
| 359 | Awareness |  |  |  |  |
| 360 | Yes | 123 | 67 | .004 | 2.570 |
| 361 | No | 20 | 23 |  |  |
| 362 |  |  |  |  |  |
| 363 | Willingness |  |  |  |  |
| 364 | Yes | 107 | 42 | .040 | 2.366 |
| 365 | No | 14 | 13 |  |  |
| 366 |  |  |  |  |  |
| 367 | Education |  |  |  |  |
| 368 | Yes | 88 | 52 | .205 | 1.405 |
| 369 | No | 53 | 44 |  |  |

## Discussion

This study demonstrates that non-participation significantly undermines schistosomiasis control efforts and sustains local transmission in the Abee community, Central Region of Ghana. Using a mixed-methods approach that combined urine microscopy, PCR, structured questionnaires, and multivariate logistic regression, the study documented a schistosomiasis prevalence of 17.1% (51/298) by microscopy and 20.1% (60/298) by PCR. Lifetime MDA participation was only 48.7%, with 31.5% of participants reporting that they had never received praziquantel. These findings confirm that Abee remains a persistent transmission hotspot despite national progress in reducing the schistosomiasis burden in Ghana.

The majority of participants were young, with 28.2% aged 0-10 years and 50.3% aged 11-20 years. This age distribution is consistent with the typical epidemiology of urogenital schistosomiasis, which predominantly affects school-aged children due to frequent contact with infested freshwater bodies. The prevalence observed in this study is considerably higher than the current national estimate of approximately 6.8% [5], yet comparable to localized findings in the Central Region, such as the 15.45% reported in the nearby Esuekyir community using similar microscopy methods [26].

The higher detection rate of PCR relative to microscopy illustrates the improved sensitivity of molecular diagnostics for identifying low-intensity and subpatent infections that are often missed by conventional parasitological methods. This diagnostic gap has practical implications, as overreliance on microscopy alone may underestimate the true human reservoir and create a misleading picture of MDA effectiveness. Multivariate analysis showed that non-participation in MDA and lack of disease education were significant independent predictors of infection. Individuals who had never received treatment exhibited higher infection rates (24.2% vs. 14.7%), while lack of education on schistosomiasis doubled the odds of infection (AOR = 2.389, p = 0.018). In contrast, awareness of the disease was strongly associated with MDA uptake (OR = 2.570, p = 0.004). These results are consistent with previous studies indicating that MDA coverage below 75% is generally insufficient to interrupt *Schistosoma haematobium* transmission, especially in areas with continued environmental contamination [14, 15]. Untreated individuals continue to excrete viable eggs into freshwater bodies, thereby maintaining the snail-human transmission cycle despite repeated annual MDA. The issue is made worse by inadequate Water, Sanitation, and Hygiene (WASH) conditions. In this study, 12.8% of participants utilized public restrooms, 43.3% shared restrooms, and a significant percentage used trash dumps or engaged in open defecation.

These inadequate sanitation methods make it easier for *S. haematobium* eggs to contaminate water bodies, maintaining high transmission intensity regardless of individual treatment coverage. The detection of microhaematuria (6.7%) and leukocyturia (8.4%) in some participants further provides clinical evidence of ongoing urogenital pathology linked to active infection. These findings align with broader literature emphasizing that integrated WASH interventions are essential alongside MDA for sustainable schistosomiasis control [12].

The strengths of this study include the use of both conventional and molecular diagnostic methods on the same samples, the combination of parasitological and behavioral data, and the application of multivariate analysis to identify independent predictors. By focusing on a single high-burden community with a predominantly young population, the study offers detailed and specific insights that complement national-level surveys.

However, some limitations must be acknowledged. The cross-sectional design limits causal inference regarding the relationship between MDA non-participation and infection status. Self-reported MDA history is susceptible to recall bias, and age data were missing for 13 participants (4.4%). In addition, the study being conducted in a single community may restrict generalizability to other settings in Ghana.

Furthermore, although PCR increased diagnostic sensitivity, amplicon sequencing was not performed to confirm species identity or detect potential hybrids. The public health implications of these findings are substantial. Low MDA participation, driven by limited awareness and weak WASH infrastructure, creates and sustains focal transmission hotspots that threaten Ghana’s progress toward the WHO 2030 elimination targets. This study reinforces the need to shift from purely vertical MDA programmes to more integrated, community-centered approaches that address both treatment coverage and environmental risk factors. Future research should adopt longitudinal designs to better assess reinfection rates and long-term MDA compliance, particularly among school-aged children who bear the highest burden. In-depth qualitative studies exploring community beliefs, myths, and trust in MDA programmes would also be valuable. Additionally, intervention studies evaluating context-specific strategies such as community-led total sanitation combined with targeted health education are warranted.

This study, therefore, seeks to provide strong evidence that non-participation in MDA, underpinned by low awareness and inadequate sanitation, sustains schistosomiasis transmission in endemic communities in Ghana. Achieving sustainable control and eventual elimination will require holistic interventions that simultaneously improve treatment coverage, health literacy, and WASH infrastructure. Only through such comprehensive strategies can the transmission cycle be interrupted and the WHO 2030 goals attained.

Non-participation in MDA, driven by low awareness and barriers to treatment access, continues to sustain schistosomiasis transmission within the study population, as reflected by prevalence estimates of 17.1% by microscopy and 20.1% by PCR. The higher prevalence detected by PCR highlights the importance of sensitive diagnostic tools for identifying infections that may be missed through conventional microscopy. These findings underscore the critical need for comprehensive MDA coverage, strengthened community engagement, targeted health education, and improved treatment accessibility. Without sustained and integrated interventions, endemic communities may continue to serve as transmission hotspots, threatening progress toward the WHO 2030 schistosomiasis elimination targets. This study demonstrates that active community participation is critical to the success of MDA programmes and to breaking the cycle of schistosomiasis transmission.

## Data Availability

All previously available sequences used are included in the tables.

## Acknowledgments

We acknowledge the study participants, community leaders, as well as School authority in the Abee Community.

## Author contributions

**Conceptualization:** George Ghartey-Kwansah, Alberta Serwah Anning,

**Data curation**: George Ghartey-Kwansah, Emmanuel Mensah, Alberta Serwah Anning, Dorcas Sekyi, Frank Agyei, Godson Sodzah, Herbert Adjei, Gloria Hagan, Michael Asare & Prince Anaglatey

**Formal analysis:** Emmanuel Mensah, Frank Agyei, Godson Sodzah, Emmanuella Akumeniwaa Nkrumah, Suzzana Dickson Buabeng

**Funding acquisition:** George Ghartey-Kwansah

**Methodology:** George Ghartey-Kwansah, Emmanuel Mensah, Alberta Serwah Anning

**Project administration:** George Ghartey-Kwansah, Emmanuel Adu Sarpong, Alberta Serwah Anning

**Software**: Emmanuel Mensah, Dorcas Sekyi, Frank Agyei, Godson Sodzah, Herbert Adjei, Gloria Hagan.

**Supervision:** George Ghartey-Kwansah, Alberta Serwah Anning

**Visualization**: Emmanuel Mensah, Dorcas Sekyi, Frank Agyei, Godson Sodzah, Herbert Adjei, Gloria Hagan.

**Writing – original draft:** Emmanuel Mensah, Emmanuella Akumeniwaa Nkrumah, Suzzana Dickson Buabeng,

**Writing – review & editing:** George Ghartey-Kwansah, Alberta Serwah Anning, Emmanuella Akumeniwaa Nkrumah, Suzzana Dickson Buabeng, Emmanuel Adu Sarpong, Nicholas Donkor,

## References

1. Lackey, Horrall. Schistosomiasis. StatPearls Publishing. 2023.

2. Wandklebson Silva da P, Malcolm SD, Amélia Ribeiro de J, Karina Conceição GMdA, Allan Dantas Dos S, Márcio B-S. Population-based, spatiotemporal modeling of social risk factors and mortality from schistosomiasis in Brazil between 1999 and 2018. Acta tropica. 2021.

3. Inobaya MT, Chau TN, Ng S-K, Macdougall C, Olveda RM, Tallo VL, et al. Mass drug administration and the sustainable control of schistosomiasis: an evaluation of treatment compliance in the rural Philippines. Parasites & Vectors. 2018;11(1). doi: 10.1186/s13071-018-3022-2.

4. Qin L, Yin-Long L, Ying-Su G, Shi-Zhen L, Qiang W, Wei-Na L, et al. Global trends of schistosomiasis burden from 1990 to 2021 across 204 countries and territories: Findings from GBD 2021 Study. Acta tropica. 2024.

5. Opare, Hervie T, Mensah E, Brown-Davies C, Asiedu O, Alomatu B, et al. Schistosomiasis in Ghana from baseline to now: the impact of fifteen years of interventions. Frontiers in Public Health. 2025;13. doi: 10.3389/fpubh.2025.1554069.

6. Yared N, Teshome B, Eshetu C, Sisay T, Dereje Oljira D, Dereje G, et al. Intestinal schistosomiasis in remote areas of Southwest Ethiopia, a target region for large-scale mass drug administration. Scientific reports. 2025.

7. Tabo Z, Wangalwa R, Rwibutso M, Breuer L, Albrecht C. Future climate and demographic changes will almost double the risk of schistosomiasis transmission in the Lake Victoria Basin. One Health. 2025;21:101148. doi: 10.1016/j.onehlt.2025.101148.

8. Rinaldi G, Neil DY, Jared DH, Paul JB, Robin BG, Michael HH. New research tools for urogenital schistosomiasis. The Journal of infectious diseases. 2015.

9. Nathan CL, Fernando Schemelzer Moraes B, Daniel GC, Fiona MF, Mamoun H, Narcis K, et al. Review of 2022 WHO guidelines on the control and elimination of schistosomiasis. The Lancet Infectious diseases. 2022.

10. Humphries D, Nguyen S, Boakye D, Wilson M, Cappello M. The promise and pitfalls of mass drug administration to control intestinal helminth infections. Current Opinion in Infectious Diseases. 2012;25(5):584–9. doi: 10.1097/qco.0b013e328357e4cf.

11. Leonardo L, Rivera P, Saniel O, Villacorte E, Lebanan MA, Crisostomo B, et al. A National Baseline Prevalence Survey of Schistosomiasis in the Philippines Using Stratified Two-Step Systematic Cluster Sampling Design. Journal of Tropical Medicine. 2012;2012:1–8. doi: 10.1155/2012/936128.

12. Boateng EM, Dvorak J, Ayi I, Chanova M. A literature review of schistosomiasis in Ghana: a reference for bridging the research and control gap. Transactions of The Royal Society of Tropical Medicine and Hygiene. 2023;117(6):407–17. doi: 10.1093/trstmh/trac134.

13. Remigio MO, Veronica T, David UO, Marianette TI, Thao NC, Allen GR. National survey data for zoonotic schistosomiasis in the Philippines grossly underestimates the true burden of disease within endemic zones: implications for future control. International journal of infectious diseases: IJID : official publication of the International Society for Infectious Diseases. 2016.

14. Gurarie D, Yoon N, Li E, Ndeffo-Mbah M, Durham D, Phillips AE, et al. Modelling control of Schistosoma haematobium infection: predictions of the long-term impact of mass drug administration in Africa. Parasites & Vectors. 2015;8(1). doi: 10.1186/s13071-015-1144-3.

15. Krentel A, Fischer PU, Weil GJ. A Review of Factors That Influence Individual Compliance with Mass Drug Administration for Elimination of Lymphatic Filariasis. PLoS Neglected Tropical Diseases. 2013;7(11):e2447. doi: 10.1371/journal.pntd.0002447.

16. Amin ASMA, Roopma W. Helminthiasis. StatPearls Publishing. 2023.

17. Nicole H, Leon C. Global Health Impact: A Model to Alleviate the Burden and Expand Access to Treatment of Neglected Tropical Diseases. The American journal of tropical medicine and hygiene. 2023.

18. WHO WHO. Accelerating work to overcome the global impact of neglected tropical diseases: A roadmap for implementation. Geneva: World Health Organization; 2012. WHO publications. 2012.

19. Yajima A, Mikhailov A, Mbabazi PS, Gabrielli AF, Minchiotti S, Montresor A, et al. Preventive Chemotherapy and Transmission Control (PCT) databank: a tool for planning, implementation and monitoring of integrated preventive chemotherapy for control of neglected tropical diseases. Transactions of the Royal Society of Tropical Medicine and Hygiene. 2012;106(4):215–22. doi: 10.1016/j.trstmh.2012.01.003.

20. Adam S, Hikabasa H, Joseph Mumba Z. How community engagement strategies shape participation in mass drug administration programmes for lymphatic filariasis: The case of Luangwa District, Zambia. PLoS neglected tropical diseases. 2019.

21. Collins SKA, Eric K, Susan A-A, Joseph O, Dziedzom Komi de S. Community perspectives on persistent transmission of lymphatic filariasis in three hotspot districts in Ghana after 15 rounds of mass drug administration: a qualitative assessment. BMC public health. 2018.

22. Tansy E, Elizabeth A, Emma MH-E, John H, Sarah EB, Martin JH, et al. Non-participation during azithromycin mass treatment for trachoma in The Gambia: heterogeneity and risk factors. PLoS neglected tropical diseases. 2014.

23. Kakoba GA, Omara D, Wananda Y, Nambatya J, Charles Okiria J, Edielu A. Determinants of Uptake of Mass Drug Administration for Schistosomiasis Control in Butiaba Sub-county, Uganda. International Journal of TROPICAL DISEASE & Health. 2023:26–42. doi: 10.9734/ijtdh/2023/v44i31393.

24. Duah E, Kenu E, Adela EM, Halm HA, Agoni C, Kumi RO. Assessment of urogenital schistosomiasis among basic school children in selected communities along major rivers in the central region of Ghana. The Pan African medical journal. 2021;40(96):1937–8688 (Electronic). doi: 10.11604/pamj.2021.40.96.26708. PubMed Central PMCID: PMCPMC8607954.

25. Rodríguez Del Á, Ar G-R. Sample size calculation. Allergologia et immunopathologia. 2014.

26. Opoku-Kwabi D, Sevor B, Sarpong EA, Sam PK, Frimpong AA, Marfo PA, et al. Prevalence of schistosomiasis among school children at Esuekyir community in the Central Region of Ghana. BMC Infectious Diseases. 2024;24(1). doi: 10.1186/s12879-024-09928-3.

27. Cnops L, Soentjens P, Clerinx J, Van Esbroeck M. A Schistosoma haematobium-Specific Real-Time PCR for Diagnosis of Urogenital Schistosomiasis in Serum Samples of International Travelers and Migrants. PLoS Neglected Tropical Diseases. 2013;7(8):e2413. doi: 10.1371/journal.pntd.0002413.

28. Schols R, Carolus H, Hammoud C, Mulero S, Mudavanhu A, Huyse T. A rapid diagnostic multiplex PCR approach for xenomonitoring of human and animal schistosomiasis in a ‘One Health’ context. Transactions of The Royal Society of Tropical Medicine and Hygiene. 2019;113(11):722–9. doi: 10.1093/trstmh/trz067.

